# Genetic relationships and biobehavioral pathways between suicidality and comorbid mental disorders: a comprehensive cross-phenotype analysis

**DOI:** 10.1101/2025.01.26.25321131

**Authors:** Yunqi Huang, Ze Meng, Shiwan Tao, Yunjia Liu, Lisa R. Miller-Matero, Esti Iturralde, Xiaoyu Liang, Albert M. Levin, Brian K. Ahmedani, Qiang Wang, Hongsheng Gui

## Abstract

**Background:** The close relationships between mental disorders and suicidality are frequently seen in epidemiology. Shared genetic liabilities and brain structure variation may underlie these associations. Therefore, we aimed to investigate the phenotypic and polygenetic associations between multiple mental disorders and different levels of suicidality, as well as mediation by grey morphology and white matter tracts.

**Methods:** Using raw data from the UK Biobank (UKB) European population, we first evaluated the phenotypic and polygenic relationships between 12 mental disorders and gradient scales of suicidality. We then accessed data from the All of Us (AoU) diverse cohort to replicate findings in European and African American populations. Demographic and social factors, including age, sex, BMI, education, deprivation, income, smoking, and chronic pain were included as covariates. Second, we used existing genome-wide association study (GWAS) summary statistics from 12 major mental conditions to estimate genetic correlations and identify pleiotropic genes using a combination of statistical genetics tools. Third, we further explored the potential mediation effects of brain structure on the relationship between mental disorders and suicidality through structural equation modeling and Mendelian randomization analyses.

**Results:** In the UKB European population, 150,861 eligible individuals were retained after standard GWAS quality control. Nine out of 12 mental disorders showed both significant phenotypic and polygenic correlations with gradient suicidality (*P*_bonferroni_ <0.05). Using GWAS summary statistics, we also observed positive global and regional genetic correlations between the 12 mental disorders and suicidality (r_g_ ranging from 0.25 to 0.68, *P*_bonferroni_ <0.05). Across pairs of suicidality and other mental disorders, we identified 73 out of 136 pleiotropic functional genes (including 58 novel ones associated with suicidality) shared by two or more pairs. These genes were enriched in pathways including regulation of immune system process and DNA, nucleosome and chromatin organization, and phenotypes as common mental disorders and brain morphology. Finally, the association between mental disorders and suicidality was significantly mediated by several structural brain imaging features.

**Conclusion:** This study underscores the urgent need to address the shared and distinct genetic architecture of suicidality and its related mental conditions. Combining longitudinal population-level biobanks with disease-ascertained GWAS data is warranted to further enhance our understanding of this complex phenomenon. Our research findings will guide future suicide prevention and precise treatment among individuals with or without major mental disorders.

## Introduction

Suicide is a major global cause of death^1^. Worldwide, suicide is the fifteenth leading cause of death, accounting for 1.4% of all mortality^2^. In addition to suicide deaths, suicidal ideation (SI)^3^ and nonfatal suicide attempts (SA)^4^ also warrant attention. Globally, lifetime prevalence rates are approximately 9.2% for SI and 2.7% for SA^5^. As a more general term, ‘suicidality’ encompasses a broad range of experiences and behaviors, from suicidal ideations, to acts of deliberate self-harm and suicide attempts, occurring along a spectrum towards completed suicide, although the progression from ideation to behavior is not always linear^6^. By any measure, there is an urgency to better understand and prevent suicide by understanding and intervening in both SI and SA.

Although there are various risk factors for suicide, including neurobiological factors, socioeconomic factors, and personality traits^7–9^, epidemiological and psychological autopsy studies have identified psychopathology as an important predictor of suicidality^10^, mostly due to comorbidity of major depressive disorder (MDD) and bipolar disorder (BD), followed by anxiety disorders (ANX), posttraumatic stress disorder (PTSD), obsessive-compulsive disorder (OCD), substance use disorders (e.g., alcohol [AUD], cannabis [CUD] and opioids [OUD]), attention deficit hyperactivity disorder (ADHD) and autism spectrum disorder (ASD), eating disorders (e.g., anorexia nervosa [AN]), and psychotic disorders such as schizophrenia (SCZ). However, previous studies could not specifically address common pathology because they either examined one diagnostic group, or had too few cases to stratify or separate diagnostic groups^11^. Therefore, further investigation is needed to understand the common pathology and etiology underlying the phenotypic linkage between suicidality and multiple common mental disorders.

According to the diathesis-stress model, genetic components and brain structural and DTI variation play an important role in the diathesis of suicidality^12^, as well as the phenotypic linkage between suicidality and mental disorders^13^. Family, twin and adoption studies suggest a heritability estimate for suicidality of approximately ranging from 30% to 55%^14–16^. And heritability for mental disorders has been estimated from 26% in PTSD^17^ up to 80% in SCZ^18^. Genome-wide association studies (GWAS) have identified significant genetic liability between mental disorders and suicidal phenotypes (SI and SA) individually^13,19–24^. However, there is a lack of consideration for how such genetic profiles align with mental disorders and gradient suicidality. Furthermore, both genetics and imaging studies have shown numerous but small associations^25,26^ with psychiatric disorders^27^ or suicidality^28^. Therefore, a comprehensive approach that combines genetics and imaging could help us understand the underlying mechanisms of the phenotypic linkage between suicidality and multiple common mental disorders.

To quantify the shared etiology underlying suicidality and multiple mental disorders, we combined UK Biobank (UKB; as discovery) and All of Us (AoU; as replication) phenome-genome data and published GWAS summary statistics (as annotation) to comprehensively analyze phenotypic linkage and polygenic overlap (polygenic risk score [PRS] estimation) between suicidality and 12 major psychiatric disorders taking into account of social environmental factors. This rigorous investigation also aimed to explore the potential mediating role of brain structures. Additionally, we sought to perform quantitative syntheses of frequently comorbid psychiatric disorders found in suicidality and to investigate the possible shared genetic architectures (horizontal pleiotropy) among pairs involving suicidality and one of those 12 psychiatric disorders. The statistical genetics approaches employed were carefully tailored to the data, enabling us to assess the associations between SA and each of the 12 psychiatric disorders, as well as their commonalities. The insights gained from this approach could inform the development of more efficient and effective strategies for managing these outcomes and promoting overall behavioral health in the future.

## Materials and Methods

### 1. Study design and Data resource

Genomic data and statistical genetics analyses provide a novel approach to disentangle the comorbidity between mental disorders and suicidality. The primary discovery was conducted using data from UKB, which recruited more than 500,000 participants aged 40–69 years from 2006 to 2010 and collected a wide range of phenotype and genotype data. A detailed description of the UKB data has been provided elsewhere^29^. A replication study was conducted in AoU^30^, launched in 2018 by the National Institutes of Health of the United States. This study has released linked phenotypic and genotype data from 312,940 participants (controlled tier v7 data released in Feb 2023)^31^.

Table 1 provides a summary of the data resource used in this study, including primary data (UKB [application No. 86920], AoU) and secondary data (GWAS summary statistics). We designed an integrated analytical pipeline to investigate their relationship (Supplementary Figure 1). This protocol was approved by Institutional Review Board (IRB) of Henry Ford Health (project No. 16896-01). The UKB received ethical approval from the North West–Haydock Research Ethics Committee (REC reference 21/NW/0157), and appropriate informed consent was obtained from all participants. All data from the AoU program was accessed on the designated Researcher Workbench cloud platform, following a stringent data usage policy for confidentiality and safety^32^.

**Table 1:** main data resources from public GWAS summary statistics and databases.

| Phenotype* | Data type | Resource | Analysis# | population | summary data source | cases | controls |
| --- | --- | --- | --- | --- | --- | --- | --- |
| SCZ/schizophrenia | GWAS Summary statistics | PGC | 2SMR, MVMR, LSDC, LAVA, PLACO | EUR/AFR | 35396580/<br>36515925 | 52017(EUR)<br>/5998(AFR) | 75889(EUR)<br>/3826(AFR) |
|  | Raw phenotypes and genotypes | UKB | Epi, PRS training, SEM, 2SMR, MVMR, LSDC, LAVA, PLACO | EUR | / | 2617 | 470059 |
|  | Raw phenotypes and genotypes | AoU | Epi, PRS training, SEM | EUR/AFR | / | 8566 | 357991 |
| BD/bipolar disorder | GWAS Summary statistics | PGC | 2SMR, MVMR, LSDC, LAVA, PLACO | EUR | 34002096 | 41,917 | 371,549 |
|  | Raw phenotypes and genotypes | UKB | Epi, PRS training, SEM, 2SMR, MVMR, LSDC, LAVA, PLACO | EUR | / | 2827 | 469849 |
|  | Raw phenotypes and genotypes | AoU | Epi, PRS training, SEM | EUR/AFR | / | 15759 | 350798 |
| MDD/major depressive disorder | GWAS Summary statistics | iPSYCH | 2SMR, MVMR, LSDC, LAVA, PLACO | EUR/AFR | 37464041/<br>34045744 | 246363<br>(EUR)/ | 561190(EUR)<br>/ |
|  | Raw phenotypes and genotypes | UKB | Epi, PRS training, SEM, 2SMR, MVMR, LSDC, LAVA, PLACO | EUR | / | 71644 | 401032 |
|  | Raw phenotypes and genotypes | AoU | Epi, PRS training, SEM | EUR/AFR | / | 95525 | 271032 |
| ASD/autism | GWAS Summary statistics | PGC | 2SMR, MVMR, LSDC, LAVA, PLACO | EUR | 30804558 | 18,381 | 27,969 |
|  | Raw phenotypes and genotypes | UKB | Epi, PRS training, SEM, 2SMR, MVMR, LSDC, LAVA, PLACO | EUR | / | 426 | 472250 |
|  | Raw phenotypes and genotypes | AoU | Epi, PRS training, SEM | EUR/AFR | / | 1410 | 365147 |
| ADHD/attention deficit hyperactivity disorder | GWAS Summary statistics | PGC | 2SMR, MVMR, LSDC, LAVA, PLACO | EUR | 36702997 | 38,691 | 186,843 |
|  | Raw phenotypes and genotypes | UKB | Epi, PRS training, SEM, 2SMR, MVMR, LSDC, LAVA, PLACO | EUR | / | 157 | 472519 |
|  | Raw phenotypes and genotypes | AoU | Epi, PRS training, SEM | EUR/AFR | / | 14822 | 351735 |
| PTSD/post-traumatic stress disorder | GWAS Summary statistics | PGC | 2SMR, MVMR, LSDC, LAVA, PLACO | EUR | 31594949 | 32,428 | 174,227 |
|  | Raw phenotypes and genotypes | UKB | Epi, PRS training, SEM, 2SMR, MVMR, LSDC, LAVA, PLACO | EUR | / | 2747 | 469929 |
|  | Raw phenotypes and genotypes | AoU | Epi, PRS training, SEM | EUR/AFR | / | 41403 | 325154 |
| AUD/alcohol use disorder | Summary statistics | MVP | 2SMR, MVMR, LSDC, LAVA, PLACO | EUR/AFR | 32451486/<br>30940813 | 57564(EUR)<br>/17267(AFR) | 256399(EUR)<br>/39381(AFR) |
|  | Raw phenotypes and genotypes | UKB | Epi, PRS training, SEM, 2SMR, MVMR, LSDC, LAVA, PLACO | EUR | / | 11014 | 461662 |
|  | Raw phenotypes and genotypes | AoU | Epi, PRS training, SEM | EUR/AFR | / | 43615 | 322942 |
| CUD/cannabis use disorder | GWAS Summary statistics | PGC | 2SMR, MVMR, LSDC, LAVA, PLACO | EUR | 33096046 | 14,080 | 343,726 |
|  | Raw phenotypes and genotypes | UKB | Epi, PRS training, SEM, 2SMR, MVMR, LSDC, LAVA, PLACO | EUR | / | 382 | 472294 |
|  | Raw phenotypes and genotypes | AoU | Epi, PRS training, SEM | EUR/AFR | / | 10646 | 355911 |
| OUD/opioid use disorder | Summary statistics | MVP | 2SMR, MVMR, LSDC, LAVA, PLACO | EUR/AFR | 32492095/<br>34488071 | 10544(EUR)<br>/ | 72163(EUR)<br>/ |
|  | Raw phenotypes and genotypes | UKB | Epi, PRS training, SEM, 2SMR, MVMR, LSDC, LAVA, PLACO | EUR | / | 262 | 472414 |
|  | Raw phenotypes and genotypes | AoU | Epi, PRS training, SEM | EUR/AFR | / | 13514 | 353043 |
| AN/anorexia nervosa | GWAS Summary statistics | PGC | 2SMR, MVMR, LSDC, LAVA, PLACO | EUR | 31308545 | 16,992 | 55,525 |
|  | Raw phenotypes and genotypes | UKB | Epi, PRS training, SEM, 2SMR, MVMR, LSDC, LAVA, PLACO | EUR | / | 2138 | 470538 |
|  | Raw phenotypes and genotypes | AoU | Epi, PRS training, SEM | EUR/AFR | / | 7418 | 359139 |
| OCD/opioid use disorder | GWAS Summary statistics | PGC | 2SMR, MVMR, LSDC, LAVA, PLACO | EUR | 28761083 | 2,688 | 7,037 |
|  | Raw phenotypes and genotypes | UKB | Epi, PRS training, SEM, 2SMR, MVMR, LSDC, LAVA, PLACO | EUR | / | 1338 | 471338 |
|  | Raw phenotypes and genotypes | AoU | Epi, PRS training, SEM | EUR/AFR | / | 1938 | 364619 |
| ANX/anxiety | Summary statistics | Meta | 2SMR, MVMR, LSDC, LAVA, PLACO | EUR/AFR | 37945807/<br>31712720/<br>31906708 | 74973(EUR)<br>/5664(AFR) | 400243(EUR)<br>/18784(AFR) |
|  | Raw phenotypes and genotypes | UKB | Epi, PRS training, SEM, 2SMR, MVMR, LSDC, LAVA, PLACO | EUR | / | 53268 | 419408 |
|  | Raw phenotypes and genotypes | AoU | Epi, PRS training, SEM | EUR/AFR | / | 88492 | 278065 |
| suicidality | Raw phenotypes and genotypes | UKB | Epi, PRS target, SEM | EUR | / | score 0=102125, score 1=26268, score 2=16139, score 3=3125, score 4=6750 |  |
|  | Raw phenotypes and genotypes | AoU | Epi, PRS target, SEM | EUR/AFR | / | score 0=14990, score 1=14990, score 2=4688 |  |
| PGCSA/suicide attempts and death | Summary statistics | PGC | MTAG | EUR | 37777856 | 43,871 | 915,025 |
| IDE/suicide ideation | Summary statistics | MVP | MTAG, PRS training | EUR | 36940203 | 62,023 | 376,826 |
| IPSYCHSA/suicide attempts | Summary statistics | iPSYCH | MTAG, PRS training | EUR | 30116032 | 6,024 | 44,240 |
| SITB/suicide ideation and attempts as a binary phenotype | Summary statistics | MVP | PRS training | AFR | 36515925 | 30,283 | 90,835 |

\*summary data without UKB samples was selected for PRS training data

In parallel with individual-level raw data, we gathered GWAS summary statistics with no sample overlaps for SI, SA, and mental disorders, to better interrogate the connection between suicidality and its comorbidities (Table 1). Samples were comprised of individuals with either European or African populations as genetic ancestry. For mediation analyses, we also gathered European ancestry GWAS summary statistics of volume^33^, surface area^34^, thickness^34^ and fractional anisotropy (FA) of white matter skeletons^35^. For suicidality specifically, we created a new GWAS summary statistics data using MTAG^36^, which combined summary data for SI and SA. Detailed quality control and harmonization of GWAS data across public data from UKB and AoU was provided in the Supplementary Methods.

### 2. Outcomes and exposure measures

2.1. For UKB individual data, the definition of suicidality phenotypes (gradient status 0-4, spanning SI [score 1-2], SA [score 3] and SA&SD [score 4]) was determined through ICD 10 code and Self-harm behavior questionnaire, while 12 major comorbid psychiatric traits were determined through clinical records and questionnaire (Supplementary methods and Table 1). Additionally, we considered brain image features as mediators and psychosocial behavioral factors including education status, index of multiple deprivation, smoking status, multiple chronic pain, loneliness, chance to confide and risk-taking behaviors as covariates. Age and sex of participants at baseline characteristics were determined at recruitment stage. Data processing and quality control details are in Supplementary methods and Table S1.

2.2. For AoU individual data, definitions of suicidality, psychiatric disorders, and potential covariates were kept the same (or as close as possible) to those assessed in UKB. Details are provided in Table S2. Then we used self-reported ancestry to select European-descent and African individuals following the same processing of genotype data quality control as in the UKB (Supplementary methods). Brain imaging data was not available as it was not included in AoU program.

### 3. Statistical methods

#### 3.1 Cross-phenotype relationship in two cohorts with individual data

##### Observational and PRS association in UKB European population

Descriptive statistics between gradient groups were compared, and tests for differences against continuous and categorial variables were conducted using independent analysis of variance (ANOVA) and chi-square statistics, respectively. Ordered logistic regression was performed to examine the observational association between mental disorders (ever or never) and suicidality outcomes (score 0-4) in all eligible UKB participants using the ‘polr’ function in the R package ‘MASS. The model was adjusted by age, gender, body mass index (BMI) and psychosocial behavioral factors. For genetic data, published GWAS were used as training datasets to develop disease-specific PRSs. The UKB data was then used as the target to calculate PRS associations. PRS-CS^37^ and PLINK^38^ were used to aggregate all weighted SNPs into one PRS for each individual in the UKB. To evaluate genetic effects, standardized PRSs were associated with ordered phenotype suicidality, adjusted for top 4 principal components (PCs)^39^, array types, and the same covariates mentioned above.

##### Sensitivity analyses

To better interpret the observational and PRS regressions better, several levels of sensitivity analyses were then conducted. First, the suicidality outcomes were categorized into binary trait: SI, SA, and SD. Second, ordered logistic regression were examined separately in males and females. Third, for deep phenotyping of exposure, we distinguished MDD with or without anxiety, MDD with or without psychotic symptoms, phobia versus panic. Fourth, longitudinal analyses were conducted focused on participants who have SI, SA or SD manifested after mental disorder diagnosis. (more details in Supplementary methods).

##### Structural equation models

Two models were used in the causal analysis: Model 1 was to examine direct and indirect effects of mental disorders on suicidality, via a path diagram specifying a direct pathway with whether mental disorders and SA PRS predicted suicidality, and indirect pathways operating conditional effects of other PRSs and psychosocial behavioral factors on mental disorders. Model 2 was to further examine potential effects of structural neuroimaging indices mediating the pathway from mental disorders towards suicidality, on condition of both genetic and psychosocial behavioral factors. Path modeling was performed using the ‘lavaan’ R package v0.6-17^40^ (Supplementary Methods).

##### Replication in AoU diverse populations

For significant association from observational, PRS association analyses and SEM model 1, the same models were performed in AoU for replicating them in European and African populations. To avoid data imbalance in exposure variables, only those mental health conditions with accessible phenotypic and genetic data in both ancestral groups were included in this replication analysis (Supplementary Methods and Table S2).

#### 3.2 Pleiotropic gene/gene-set identification with GWAS summary data

##### Pleiotropy identification and polygenic enrichment analyses

First, we utilized linkage disequilibrium score regression (LDSC)^41^ and local analysis of (co)variant annotation (LAVA, 0.1.0)^42^ for estimating genetic correlations for each pair consisting of suicidality and one of those mental disorders at a genome-wide and local (2495 regions) level, respectively. Next, to determine polygenic enrichment between mental disorder and suicidality phenotype pairs, shared loci between phenotype pairs were identified using PLACO^43^. SNPs with *P* < 5×10^−8^ after PLACO analysis were considered significant pleiotropic variants. Regions with these variants were further fine-mapped using CAVIAR^44^ implemented in Functional mapping and annotation of genetic associations (FUMA)^45^. Finally, a Bayesian colocalization analysis^46^ was performed to further identify shared causal variants in each pleiotropic locus. We declared a colocalized locus with posterior probability of H4 (PP.H4) larger than 0.7.

##### Gene prioritization and functional annotations

Based on PLACO results, we further explored the biological mechanisms of these pleiotropic loci. Gene-level multimarker analysis of GenoMic annotation (MAGMA)^47^ was applied to aggregate a set of SNP-level associations into a single gene-level association signal using summary statistics derived by PLACO. They were further validated by expression quantitative trait loci [eQTL]–informed MAGMA (e-MAGMA)^48^ and Hi-C–coupled MAGMA (h-MAGMA)^49^ to indicate the tissue and cell-type specificity. All the results were corrected using the Bonferroni method, and the significant pleiotropic genes were categorized into pair-shared (shared by ≥ 2 mental disorder-suicidality pairs) and pair-specific groups. Next, we prioritized these genes by summary-data-based Mendelian randomization (SMR)^50^ analyses, single-cell type from database PanglaoDB and CellMarker 2.0, and protein expression level in UKB OLINK data. Gene set enrichment analyses for the pleiotropic genes through FUMA gen2fun portal. Significantly enriched pathways were declared with normalized enrichment scores greater than 2 and adjusted *P* < 0.05. Protein-protein interaction (PPI) networks were constructed by STRING^51^. More analyzing details were included in the Supplementary Methods.

##### Genetic causality and mediation

To evaluate potential causal relationships (vertical pleiotropy), two-sample MR (2SMR) and multivariable MR (MVMR) analyses were performed using GWAS summary statistics from suicidality, its correlated psychiatric traits, and potential mediating brain imaging indices. Additional quality control and harmonization steps were included in the Supplementary Methods. All statistical analyses were performed in R version 4.3.2.

## Results

### The phenotypic and genotypic linkage between suicidality and mental disorders

Of the 154,407 unrelated UKB individuals included in the association with suicidality, 56.45% were female, and their mean age was 56.81 (7.60 as standard deviation [SD]) years. The characteristics of the participants are displayed in Table S3. After tabulating each of those 12 mental disorders and the gradient phenotype of suicidality in UKB, we observed significant differences in proportions for all 12 mental disorders across ordinal phenotype (Odds Ratios (OR) ranging from 2.27 for ANX to 9.52 for BD, *P* < 4.17 × 10^−3^, passing Bonferroni correction; Figure 1 and Table S4). Furthermore, when examining the effects of mental disorder PRSs on suicidality, we found significant associations for 9 out of 12 (MDD, BD, SCZ, AUD, ANX, OCD, AN, ASD and ADHD) along with two suicide-specific PRSs (SA-PRS and SI-PRS) after Bonferroni correction (OR ranging from 1.02 for OCD to 1.22 for MDD, *P* < 4.17 × 10^−3^; Fig. 1 and Table S4).

**Figure 1.**
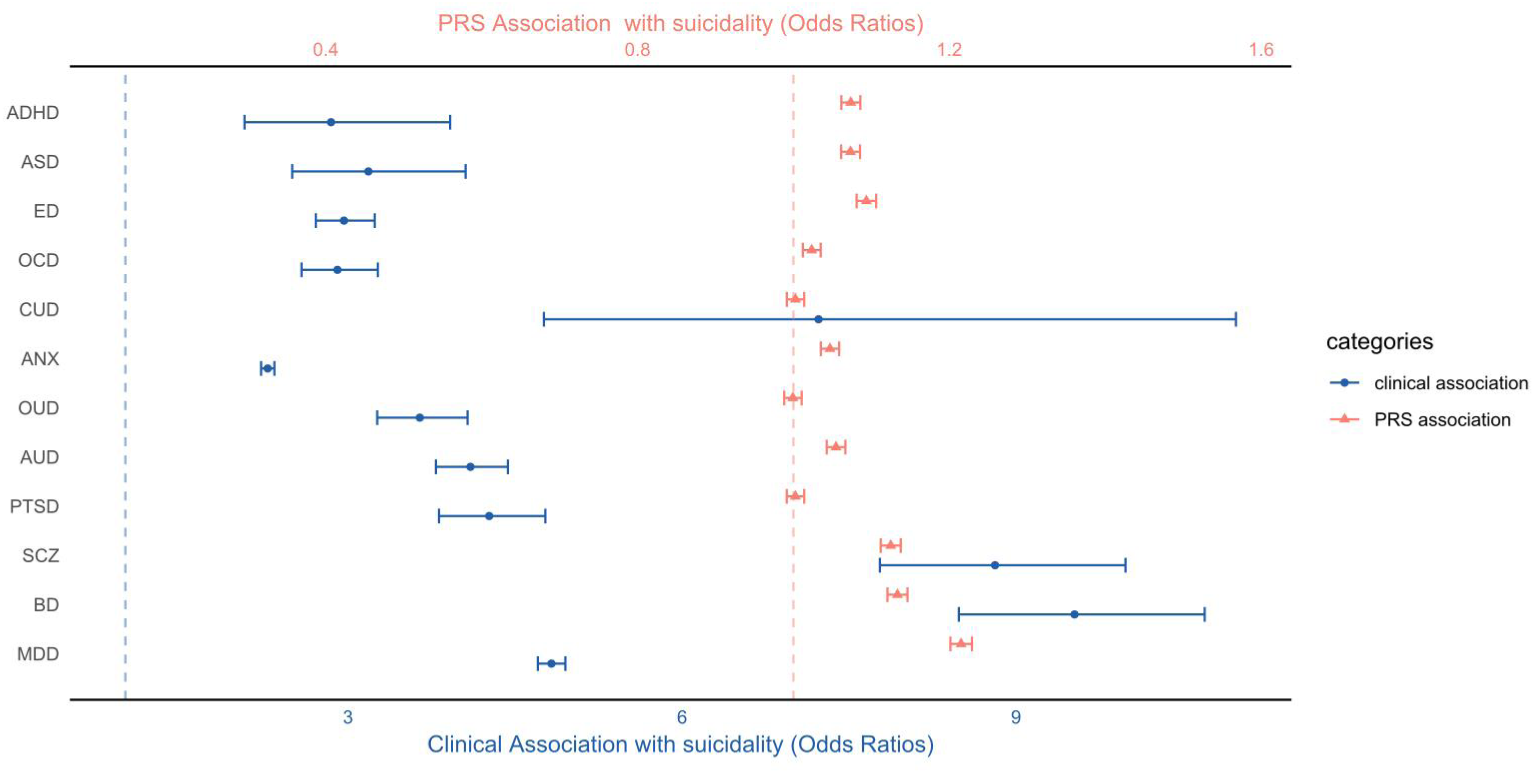
Association between 12 mental disorders and suicidality in UKB. Blue indicates clinical, while red represents mental disorder – specific associations. Both blue and red dashed lines indicate an odds ratio (OR) of 1. PRS: polygenic risk score; SA: suicidal attempts; SD: suicidal death; SCZ: schizophrenia; BD: bipolar disorder; MDD: major depressive disorder; ANX: anxiety disorder; OCD: obsessive-compulsive disorder; PTSD: posttraumatic stress disorder; ED: eating disorder; ASD: autism spectrum disorder; ADHD: Attention Deficit Hyperactivity Disorder; AUD: alcohol use disorder; CUD: cannabis use disorder; OUD: opioids use disorder.

After performing a sensitivity check for different factors, the increased risk of suicidality in patients with mental disorders remained statistically significant in both sexes, with similar effects observed for disease-specific PRSs. When zooming into different subtypes of outcome (e.g.., SI vs SA or SA vs SD), the effect sizes of each psychiatric disorder and its corresponding PRS were higher in the SA subgroup compared to the SI subgroup (e.g., OR increased from 4.68 to12.58 for MDD and from 1.20 to 1.48 for MDD-PRS); in addition two new PRS associations were found significant for SA subgroup only (OR_OUDPRS-SA_ = 1.04, 95%CI: 1.06-1.14; OR_CUDPRS-SA_ = 1.02, 95%CI: 1.00-1.05). With regard to SA vs SD comparison, only a fraction of those associations from mental disorders displayed higher OR with SD than that with SA (e.g., OR changed from 22.68 to 27.39 for SCZ and from 1.27 to 1.40 for SCZ-PRS). When further dissecting heterogeneous mental disorders (e.g., MDD and ANX) into different subtypes, a few differences were also observed (e.g., OR from 1.32 to 5.01 for MDD with or without anxiety or psychotic symptoms on suicidality). Lastly, in the longitudinal analysis, non-suicidal patients with MDD, ANX, ASD and AUD at baseline level had a significantly higher risk of manifesting suicidality in follow-up period (e.g., hazard ratio [HR]_MDD-suicidality_= 1.37, 95% CI: 1.25-1.50; HR_ANX-suicidality_ = 1.12, 95% CI: 1.05-1.20). The full results for all sensitivity analyses were shown in Supplementary Results, Figure S2 and Table S5.

After harmonizing clinical and genetic data across ancestries, five mental disorders (MDD, AUD, SCZ, ANX, and OUD) can be accessed in AoU for replication analysis. In European Americans, we validate that all of them showed significant observed and genetic correlation with suicidality (e.g., OR_MDD-suicidality_ = 1.17; OR_MDD-PRS-suicidality_= 1.25, *P* < 0.001, details in Table S4). In African Americans, we also validated all those clinical associations (ORs >1.2, *P values* <0.001); however, only PRS for schizophrenia was significantly higher in suicidal patients after Bonferroni correction (OR _SCZ-PRS-suicidality_= 1.05, 95%CI: 1.02-1.09; Fig. S3 and Table S4).

SEMs were then applied to interrogate potential causal pathways from mental disorders to suicidality. As expected, overall affection status (i.e., latent factor) of 12 mental disorders significantly contributed to higher incidence of suicidality (β = 1.69, p < 0.001; Fig. 2a) after controlling for confounding or colliding effects from suicide PRS (β = 0.008, p = 0.115), mental disorder PRSs (effect on mental phenotypes: β = 0.086, p < 0.001; effect on suicidality: β = 0.042, p = 0.001), and environmental factors (effect on mental phenotypes: β = 1.533, p < 0.001; effect on suicidality: β = 0.983, p = 0.001). Similar direction of effects was also replicated in AoU diverse cohorts, though the magnitude was smaller when using approximates from 5 mental disorders only (European or African populations; Fig. S4, Table S6).

**Figure 2.**
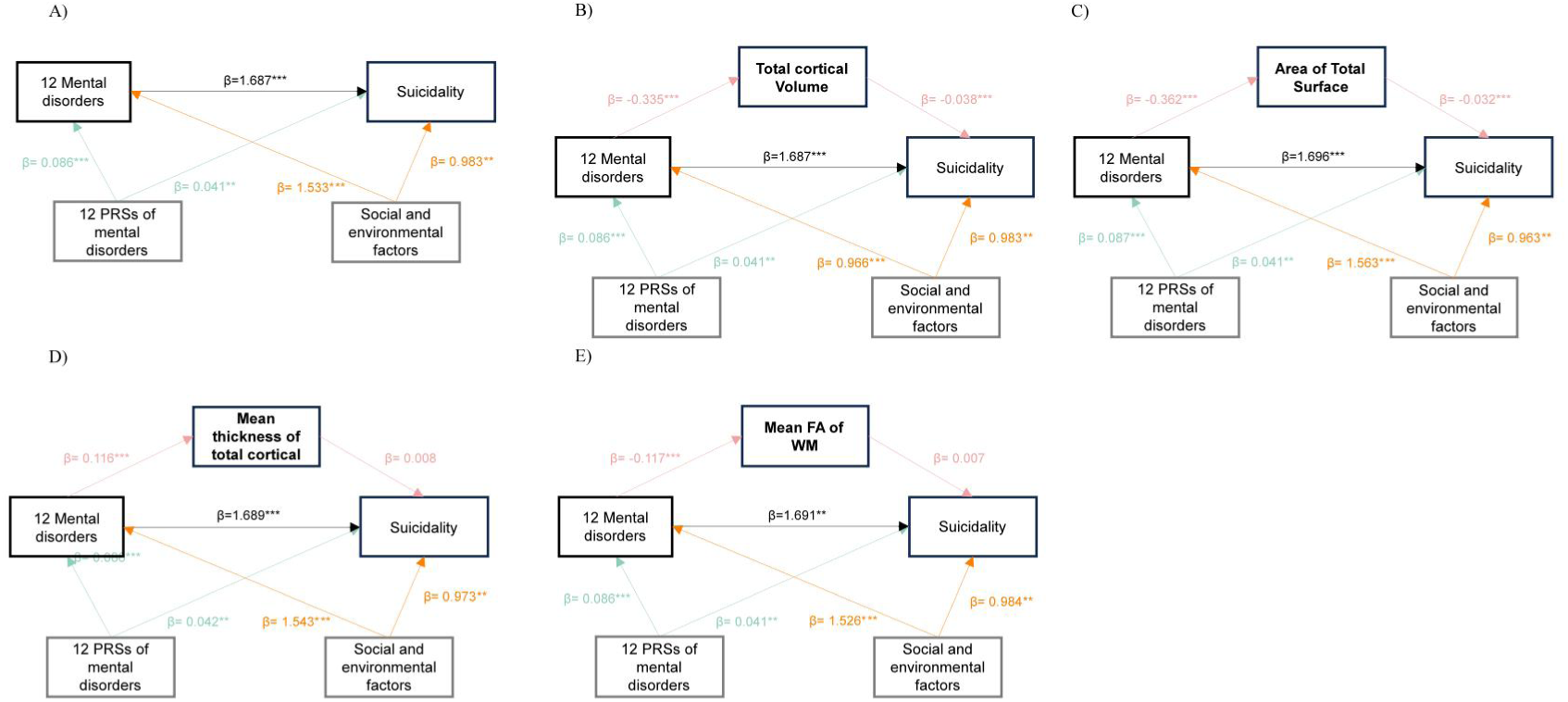
A) Structural equational models for 12 mental disorder, PRS, social environmental factors and suicidality in UK Biobank. Lifetime smoking, multi chronic pain, loneliness, confide, risk-taking, educational status, deprivation index, and BMI are as latent variables for social and environmental factors. *P < 0.05, **P < 0.01, ***P < 0.001. B-E): Based on the model in A), each brain index was added in the model and total cortical volume and area of total surface showed mediation effects between overall mental disorders and suicidality. *P < 0.05, **P < 0.01, ***P < 0.001

### The genetic correlation and causality between mental disorders and suicidality

After deriving new suicidality GWAS data by MTAG analysis, lead SNPs and risk loci increased to 16 and 13 respectively (e.g., rs7746199, *P* = 9.39 × 10^-14^; more details in supplementary Figure S5-6 and Table S7). When paired with GWAS data for those 12 mental disorders (Figure 3A and Table S8), significant positive global genetic correlations were observed for most of the trait pairs (r_g_ ranging from 0.25 with ASD to 0.68 with MDD, *P* < 4.17 × 10^−3^; r_g_ = 0.14 with OCD was nominally correlated). When zooming into specific genomic regions by LAVA analysis, all of mental disorder – suicidality pairs showed consistent local genetic correlations, and 994 out of 2495 regions remained after stringent Bonferroni correction among 11 pairs (p < 1.67 × 10^-6^), with 396 regions displaying significantly correlated among ≥2 trait pairs (Fig. 3B and Table S9). When testing putative mediation pathways using multiple GWAS data, 2SMR analyses revealed 10 significant positive associations when investigating mental disorders and suicidality separately (Table S10).

**Figure 3.**
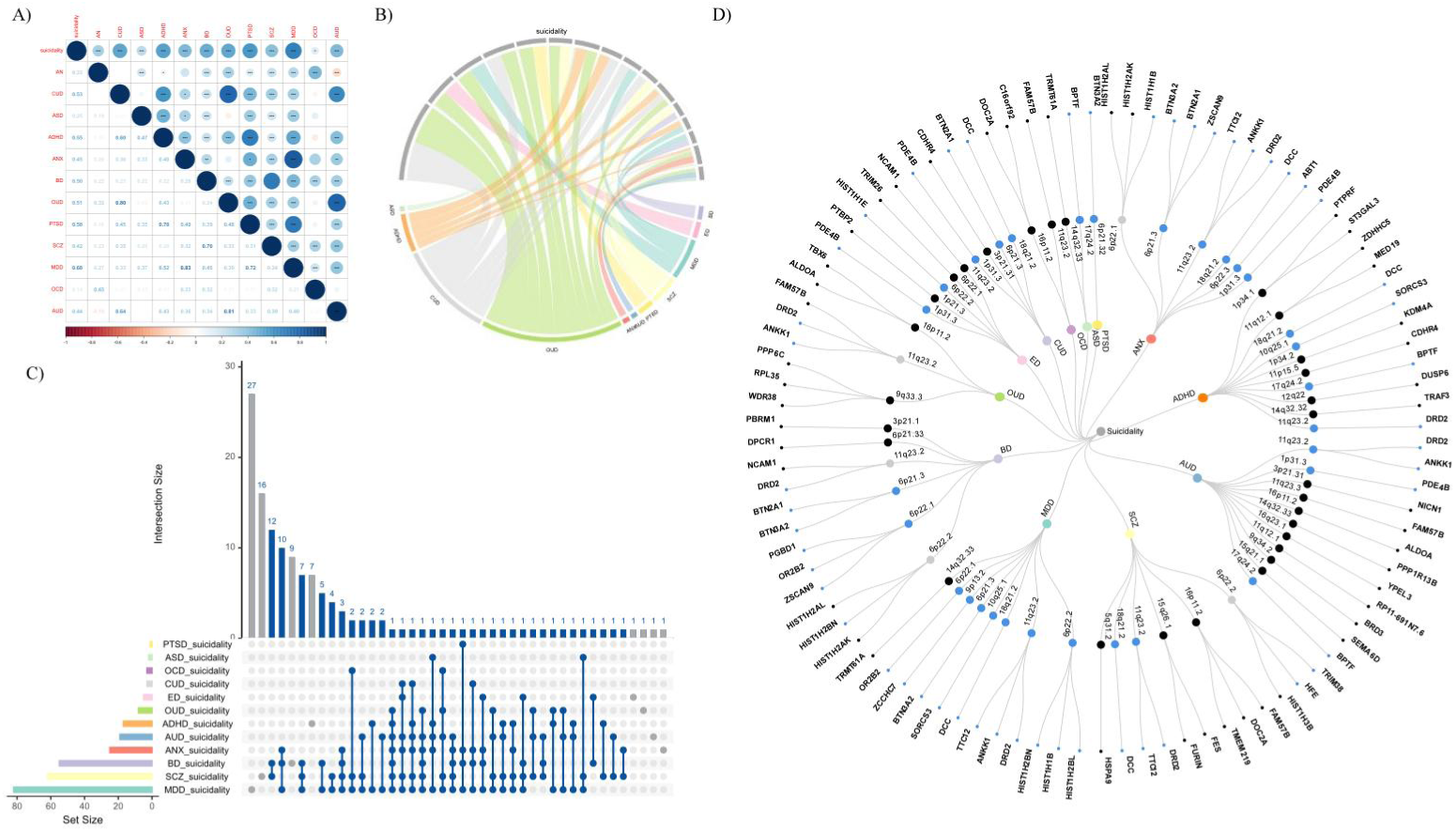
A) LDSC results between among each 12 mental disorders and suicidality. B) LAVA results between pairs of mental disorders and suicidality, and 863 out of 2495 regions displayed significantly correlated among ≥2 trait pairs. 9 pairs showed significant correlation in locus CHR3:71223282:72334704. C): 136 pleiotropic genes were finally validated by MAGMA, e-MAGMA and h-MAGMA, respect to those 73 genes shared by ≥ 2 trait pairs. D): A circular dendrogram included 12 mental disorder diseases and suicidality, resulting in 12 trait pairs. We selected top 10 genes for each pair and displayed which loci they were in. Blue dots indicated validated genes by e-MAGMA or h-MAGMA.

### Shared genetic loci and mapped genes between suicidality and mental disorders

After systematically searching for pleiotropic variants between suicidality and mental disorders, a total of 274 SNPs (238 unique), located in 233 independent genomic loci from 110 unique chromosomal cytoband regions, were identified as potential candidates involved in 11 different trait pairs (Table S11). Some pleiotropic regions were repeatedly identified in multiple pairs (e.g., 6p21.32 indexed by rs1796518, 11q24.2 indexed by rs4936277, and 18q21.32 indexed by rs8084351), suggesting the extensive multi-trait genetic overlap existed. However, most of the 238 index SNPs were located in non-genic regions (intronic or intergenic; supplementary Figure S7), with only 7 of them from exonic region of a functional gene or noncoding RNA (e.g., *DRD2*, *OPRM1*, and *LRP8;* more annotations in Table S11). This emphasized the necessity of re-mapping these pleiotropic SNPs to other functional genes with integrative approaches. Further colocalization analysis identified 38 of 100 potential pleiotropic loci with PP.H4 larger than 0.7 (Table S11). The 11q23.2 locus, which was identified as pleiotropic loci for multiple trait pairs, was also colocalized among 4 pairs. In addition, 18 pleiotropic loci were identified with PP.H3 larger than 0.70, indicating there might be more causal variants in these loci (Table S11).

### Prioritization of Candidate Pleiotropic Genes and Characterization of Phenotype and Tissue Specificity

Then, multiple gene-based methods were applied to achieve that goal. When conducting MAGMA, 178 unique and significant pleiotropic genes were mapped by those identified loci; among them, 94 genes were shared between suicidality and ≥2 mental disorders (Table S12). After cross-linking to function evidence at transcriptome (via e-MAGMA, Table S13) and epigenome (via h-MAGMA, Table S14) levels, 136 pleiotropic genes were finally validated with respect to those 73 genes shared by ≥ 2 trait pairs (Table S15 and Fig. 3C-3D), 35 were identified as novel genes to suicidal or mental disorders, along with several key genes (e.g., *DRD2 shared by 7 pairs, ANKK1, DCC, and PDE4B shared by 6 pairs*), full list in supplementary result and Table S15.

Among these 73 multi-pair shared genes, additional TWAS (via SMR) showed that expressions of 32 genes in whole blood and 13 brain tissues were associated with some of those 12 mental disorders or suicidality (e.g., *BTN3A2* and *ZCCHC7*; more in Table S15 and S16). Additional tissue (or cell) specificity analysis revealed these 73 genes were not only expressed in brain tissues but also in muscle-skeletal and heart tissue, and they were enriched in 8 post-conception-week brain samples (Fig 4A, Tables S15 and S17); moreover, expressions of 47 genes showed enrichment in multiple sub-types of neuronal cells, endothelial cells, glial cells, and microglia from human or mouse data (Table S15). Last, plasma protein analysis showed expressions for 6 proteins (BTN2A1, BTN3A2, DDR1, MAD1L1, FURIN, and NCAM1) were significantly associated with suicidality in UKB samples (Table S15 and S18).

**Figure 4.**
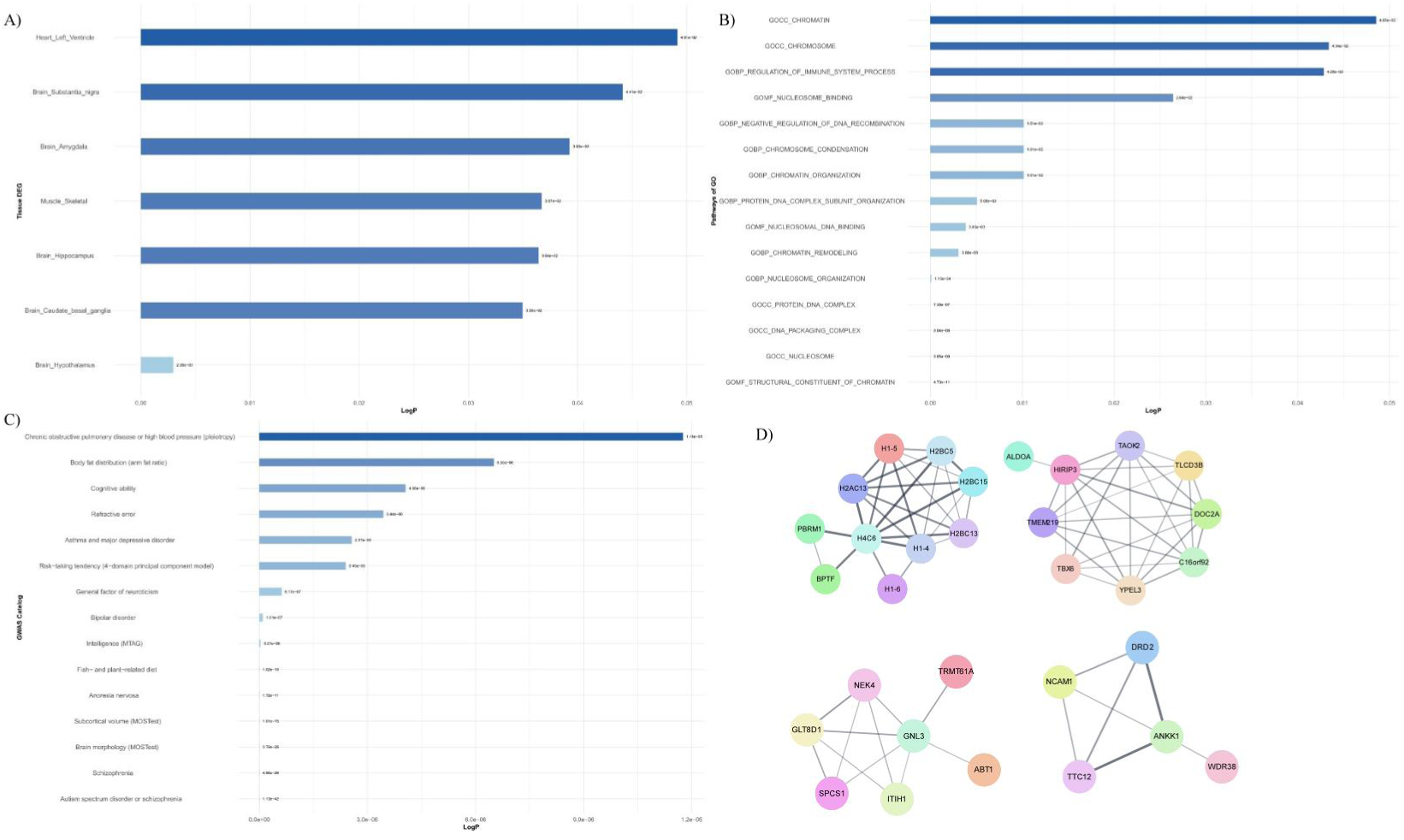
A) 7 significantly enriched tissues were identified when using the Genotype-Tissue Expression project (GTEx V8) reference panel. 139 over-represented gene-sets or pathways were firstly identified. These enriched pathways are mainly involved in regulation of immune system process and DNA, nucleosome and chromatin organization (B). Secondly, these shared genes were mainly enriched in common mental disorders, brain morphology, and behavioral traits (intelligence and risk-taking tendency) based on the latest GWAS catalog (C). PPI network analysis showed that 4 sets out of 73 multi-shared proteins form a tightly interconnected network (D).

### Enriched biological pathways and shared risk factors between suicidality and mental disorders

When grouping all 73 multi-pair shared genes for function enrichment analyses, 139 over-represented gene sets or pathways were first identified (Table S19). These enriched pathways are mainly involved in the regulation of immune system process and DNA, nucleosome and chromatin organization (Fig 4B). Secondly, these shared genes were mainly enriched in common mental disorders, brain morphology, and behavioral traits (intelligence and risk-taking tendency) based on the latest GWAS catalog (Fig. 4C). Thirdly, PPI network analysis showed that 4 sets out of 73 multi-shared proteins form a tightly interconnected network (Fig. 4D and Table S20). Lastly, extending analyses for all 136 genes suggested the co-existence of shared and distinct mechanisms when connecting suicidality and different mental disorders (supplementary Figure S8-10 and Table S21-S22).

### Causal and mediating Effects of mental disorders on suicidality

When evaluating mediating effects of neural phenotypes, the total volume of grey matters and area of total surface out of whole-brain and region-wise indices showed the most significant mediation effect between overall mental disorders and suicidality (0.75% and 0.49% mediated by total volume of grey matters and area of total surface, respectively; Fig. 2b-c, and Table S6). When examining individual pathways from each mental disorder to suicidality, various region-wise indices were instead found with the largest mediating effects accordingly (e.g., integrity of splenium of corpus callosum for SCZ-suicidality). Among them, integrity of body of corpus callosum, area of precentral, and mean thickness of postcentral were the most frequent mediators across pathways for different mental disorders (more examples in supplementary result, Figure S11, and Table S23).

MVMR analyses showed genetic diathesis for AUD and ANX were still potentially causal on suicidality after controlling for neural GWAS data. However, we did not validate significant mediation effects of brain structural indices on the causal effects from mental disorders to suicidality (Table S24).

## Discussion

In this multi-angulated study, we systematically investigated clinical and genetic relationships between suicidality and comorbid mental disorders in two population-based cohorts and multiple large-scale disease-specific GWAS datasets. Two potential types of mechanisms have been explored for their intrinsic connection: brain imaging mediators and shared susceptibility genes. Comprehensive analyses for the latter mechanism highlighted pleiotropic genetic variants, loci and genes, suggesting the involvement of immune and chromosome-related pathways. All these findings supported the shared biobehavioral pathways (via neural development and genetic liability) underlying multiple mental disorders and suicidal ideation and behaviors. Further multiple global- and region-wise brain structures have been identified as modifiable mediation factors for the former etiology mechanism.

Leveraging individual data from two cohorts, our results revealed risk of suicidality was higher in psychiatric patients or those with higher genetic diathesis (via PRS) for mental disorders, consistent with previous epidemiological evidence^10,19,20,52^. Nevertheless, clinical and genetic associations are not always consistent across traits (or subtypes), cohorts, or ancestral groups. Multiple reasons may be involved behind the observations. It is possible that the magnitude of associations may differ when the limited sample size of particular mental disorders in UKB or AoU was accessed. Also, it may be ascribed to different demographic characteristics between PRS training data (e.g., MVP consisting mainly of males) and target cohort^53,54^. Clinical association coupled with the shared genetic overlap between suicidality and psychiatric disorders, underscores the salience of mental health as a key risk category of suicide.

Moreover, regarding the 3 levels of suicidality (SI vs SA vs SD), most mental disorders didn’t display a higher incidence of SD than SA as expected, consistent with PRS association results. It might be because SD exhibited lower liability than SA^19,20^, and higher heterogeneity which should be more interpreted by gene–environment interactions^55,56^. Thus, when looking into covariable factors, we noted that female is a risk factor for SI or SA but a protective factor for SD^57,58^; education level is a risk factor for SA but a protective factor for SI. Therefore, further genetic studies focusing on deep-phenotype stratification and lifetime longitudinal data of SD, particularly considering diverse SA or SI histories, are warranted.

When focusing on post-GWAS analyses, pleiotropic genes between suicidality and psychiatric disorders were extensively distributed, with several loci especially highlighted between certain trait pairs, such as 6p21.3 (*BTN3A2, BTN2A1, ZSCAN9*), 11q23 (*ANKK1, DRD2*), 17q24.2 (*BDTF*) and 18q21.2 (*DCC*). The pleiotropic genes were more likely to be enriched in behavioral, neurological, and neurodevelopmental phenotypes, as well as specific tissues and cell types, especially brain tissues and nerve cells. In addition to being targets for antipsychotics, antidepressants, and dopamine regulators, these genes are also involved as targets for trace element supplementation, metabolism regulators, and anti-inflammatory treatment. The operation of these genes was also confirmed by the results of gene-set PRS. Additionally, functional enrichment analysis and PPI networks suggested the potential involvement of shared genetic determinant in nucleosome binding and organization, further affecting the occurrence of neuron development^59^ and immune regulation, a common pathology of mental disease^60–63^. Both DrugBank and enrichment results suggested adjunctive therapeutic options for suicidality comorbid with psychiatric disorders.

Our study also uncovered lower grey volumes, surface area, thickness and integrity of white matter tracts in several key brain regions mediated the relationships between overall status of mental health (or each mental disease) and suicidality. Notably, these findings are largely consistent with previous studies that highlighted the neurobiological substrates that total intracranial volume and total surface area showed lower trends in different levels of suicidality^64–66^. Among these regions and tracts mediating multiple trait pairs and exhibiting the most significant effects, white matter modifications of corpus callosum have been reported in several mental disorders and with history of SA and SI^67^. Additionally, precentral region was consistent with previous findings reporting hypogyrification in adult suicide attempters across corresponding mental disorders^68,69^. However, this is the first time that we discovered a role for these morphological changes in mental disorders and suicidality.

Encompassing epidemiological, genetic, and neuroimaging perspectives, this study strongly supports the Research Domain Criteria (RDoC) framework’s conceptualization of suicidality occurring across a spectrum of psychiatric conditions. Transdiagnostic and multi-omics research holds great promise for enhancing our understanding of the development of suicide^70,71^. This multidimensional approach allows for a deeper understanding of how suicidality manifests within different psychiatric conditions, facilitating the identification of shared risk factors and novel treatment targets not only in suicidality, but also in cross-disease level, ultimately improving prediction and prevention efforts in the future.

Some limitations should be considered when interpreting our findings. First, when assessing the associations between suicidality and mental disorders in the UK Biobank, though we have included potential confounding factors in our analysis, residual confounding may still exist, which may result in an overestimation of clinical associations. Second, the PRS association and pleiotropy genetic analyses may lack power due to inadequate sample size, though we applied the GWAS data sets with the largest sample size to date for each phenotype assessed, importantly for the understudied condition OCD. Further validation studies of the present findings using larger GWAS datasets are required. Finally, we were unable to assess the causal mediation effects of brain structural indices on psychiatric disorders and suicidality through mendelian randomization analyses due to insufficient power of corresponding GWAS and lack of consideration for environmental factor^72^. Advanced longitudinal data need to be further incorporated for temporal relationship.

## Conclusions

Overall, our study indicated suicidality and comorbid mental disorders present strong phenotypic and genetic association and mediation of brain structures, supported by pleiotropic genetic variants, loci, and genes with tissue and cell specificity. The shared genes were involved in immune system and chromosome formation, shared biological mechanisms concerning neurodevelopment and immune response were identified. Our findings not only support the shared genetic basis underlying multiple traits but provide novel insight into the intervention and treatment targets of mental disorders and suicidal ideation and behaviors.

## Data Availability

All data used in the present study are available online at the UK Biobank data Access Management System (https://www.ukbiobank.ac.uk/enable-your-research/apply-for-access) and the researcher workbench of the All of Us Research Program located at https://workbench.researchallofus.org

