## Supplementary files for "Genetic relationships and biobehavioral pathways between suicidality and comorbid mental disorders: a comprehensive cross-phenotype analysis"

**Huang et al.**

Supplementary methods

Supplementary results

Supplementary figures

Supplementary references

**Supplementary methods**

Here we provide additional description of data and approaches included in our analytical workflow (supplementary **Figure 1**), which was designed for mining of multiple collected datasets in this study (Table 1 and Table **S1-S2**).

**1. Biobank data processing and analytical details**

*1.1 Assessment of outcomes and exposures*

Suicidality: In UK Biobank (UKB), suicidality cases were defined by Self-harm behavior questionnaire as: 1) score 0, never thought or contemplated suicide; 2) score 1, SI, ever thought that life not worth living; 3) score 2, SI, ever contemplated self-harm; 4) score 3, SA, ever self-harmed; 5) score 4, SA or SD, ever attempted suicide or suicide death, patient-reported answers (data field 20002, 20483, and 40001), ICD 10 diagnostic codes (primary and secondary) from in-hospital records and read v2/3 codes in primary care records. Controls were selected from those Mental Health Questionnaire (MHQ) participants with answer "No" to self-harm (data field 20480); in addition, those who overlapped with any suicidality cases were also excluded. While in All of Us (AoU) Research Program, suicidality was defined by ICD 9, ICD 10, SNOMEDCode, PHQ 9 questionnaire, and an emotional health survey.

Mental disorders: cases were defined using either primary and secondary International Classification of Diseases 10th Revision (ICD 10) diagnoses codes in UKB hospital record, mental health problems ever diagnosed by a professional (electronic health records in UKB), and self-reported diagnosis mental health questionnaire (MHQ). To extract matched controls, the eligible participants need to satisfy conditions as not diagnosed with or self-reported mental disorders. In AoU, cases of mental disorders were also defined by ICD 10 and self-reported diagnosis. The detailed diagnostic codes (ICD-10, slef-report and electronic health records) and data fields in MHQ are shown in **Table S1**.

*1.2 Genotype data and quality control*

For genotype data from UKB, standard quality assessment and quality control (QA/QC) steps were applied to raw genomic data that covers all participants with SNPs directly genotyped on two Axiom arrays (~50,000 by UK BiLEVE Axiom array and ~450,000 by UK Biobank Axiom array), and SNPs imputed against Haplotype Reference Consortium reference panel (data field 22828).[1,2] To generate clean dataset for downstream association analysis, we used quality metrics provided by UK Biobank (data field category 100313 that covers genetic ethnic grouping and principal components) and those metrics (e.g., kinship relationship and genotyping quality) generated by KING or PLINK 1.9.[3,4] In general, we adopted following QA/QC steps from both sample and variant aspects.

Sample-level: 154,878 participants with suicidality phenotype were first selected by suicidality (data field 20479, 20480,20483,20485, 40001, 40002 and 41201). We filtered samples by their self-reported ancestry/ethnicity (data field 21000) to include British White (n=154,407 for observed association); and then applied category 100313 metrics to include European-ancestry individuals without sex mismatch or low genotyping individuals; lastly, we kept only unrelated samples defined by KING for polygenic risk score (PRS) analysis (n=150,861).

Variant-level: PLINK was used to filter out variants by minor allele frequency (0.01), Hardy-Weinberg equilibrium test ($1x10^{-6}$), genotyping rate (0.95), and imputation info score (0.5). This resulted in ~10M variants. A subset of common SNPs (n= 1133,960) that match with Hapmap3 variants (rsID) were retained for generating polygenic risk scores.

For genotype data from AoU, standard quality assessment and quality control (QA/QC) steps were the same to pipeline of UKB.

*1.3 Social environmental covariable selection*

Next, psychosocial and behavioral factors, like education, deprivation index, loneliness, whether able to confide, risk taking, chronic pain and whether smoke, were chosen to be covariables. Age and sex of participants at baseline characteristics were determined at recruitment stage. Data fields in psychosocial and environmental factors were shown in Table S2b. Continuous data was standardized before analysis. Noticeable, multiple chronic pain was defined as a binary phenotype by ICD 10 diagnosis, medication prescription for pain relief and whether experienced multiple pain for more than 3 months by surveys.

*1.4 outcomes selection for longitudinal studies*

The longitudinal outcomes included mental disorders, SA and SD identified by ICD-10 codes linked to the diagnosis, followed up to 28 March 2023. The endpoint for each participant was censored at the date of outcome diagnosis, the date of death or 28 March 2023, which ever occurred first. Date of mental disorder or suicidality phenotype diagnosis were determined by field of Date of first in-patient ICD-10. Participants with the following criteria were excluded: 1) participants with the following mental disorders after SI, SA or SD; 2) participants with missing data on the covariates.

The cumulative incidence of suicidality was calculated using Kaplan–Meier method implanted in 'survival' R package (3.5.7), and the log-rank test was used to determine the difference between people with or without each mental disorder. Cox proportional hazards regression models were applied to evaluate the associations between clinically or genetically associated mental disorders and suicidality phenotype.

*1.5 Neuroimaging mediator selection*

In T1 structural magnetic resonance imaging (MRI), we selected grey matter and subcutaneous nucleus volumes, surface areas and thickness, including 68 fields (33 brain regions generated with Freesurfer by parcellation of the white surface using Desikan-Killiany parcellation and 1 total index for both left and right hemisphere), respectively. In Diffusion Tensor Imaging (DTI) MRI, we selected fractional anisotropy (FA) for 27 tracts paralleled by JHU atlas each and we generated an average FA for all the skeletons. Each brain index was averaged on left and right hemispheres to match phenotypes in GWAS summary data. All data was standardized before analysis.

### 2. GWAS Data harmonization

For all the disorder GWAS summary statistics, we obtained their European-only and African-only summary statistics and performed stringent quality control: (i) excluded non-biallelic SNPs and those with strand-ambiguous alleles; (ii) SNPs that had no rs label or chromosome positions were converted by ANNOVAR[5] using Hg19 build reference avsnp150 ; (iii) SNPs without minor allele frequency (MAF) were matched by 1000 genome reference and kept SNPs that had MAF > 0.01; while SNPs whose A1 were not matched to effect allele (EA) column, we swapped A1 and A2 columns.

Next, we extracted all necessary columns (chromosome, position, rsID, alleles, sample size, odds ratio [OR], standard error [SE], and *P*). For those without OR/SE, we computed them by Z-score, sample size, *P*, and minor allele frequency (MAF).[6] Pairwise datasets were harmonized at both sample level and variant level. For PRS, both training and target cohorts were comprised of European-descent, non-overlapping, and unrelated samples. Only common SNPs (MAF >0.05) matched with HapMap3 reference were included. Variants were harmonized for their rsID or genomic coordinate, strands, and effect alleles. Variants were also handled to avoid palindrome, with genotypes from 1000 Genome CEU population[7] as reference. After QC, GWAS summary statistics were performed in PRS training and series of post-GWAS analyses.

### 3. Technical details for Sensitivity analyses and structural equation models.

For the two deep phenotypes of MDD, cases were defined by being diagnosed both MDD and

ANX/SCZ, while controls were defined by MDD patients without anxiety/psychotic symptom. PRS base data was derived from CC-GWAS[8] result (case 1:MDD with ANX/SCZ from MTAG[9] results for both GWAS summary data and case 2: MDD without ANX/SCZ from mtCOJO[10] results). For other deep phenotypes of ANX and ED, phobia versus panic, and AN versus other ED were only used in observed regression due to lacking for GWAS summary data.

In the SEM analysis, prior to hypothesis testing, all data were examined to determine missingness, identify extreme values, and confirm that the data structure meet analytic assumptions. Two models were used in the analysis: Model 1 was to examine direct and indirect effects of mental disorders on suicidality, a path model was tested specifying a direct pathway with whether mental disorders and SA PRS predicting suicidality, indirect pathways operating through PRSs of mental disorder mediated by diagnoses; Model 2 was to examine mediating effects of structural neuroimaging indices from mental disorders on suicidality. Because the chi-square test of model fit is highly sensitive to minor sources of misfit between estimated models and observed data, model fit was evaluated by meeting each alternative fit statistics, including comparative fit index (CFI; 0.95 or above indicative of good fit), root mean square error of approximation (RMSEA; 0.05 or below indicative of good fit), and standardized root mean square residual (SRMR; 0.08 or below indicative of good fit). Results were corrected by bootstrapping. Path modeling and multiple comparison were performed in R version 4.3.2 using the lavaan[11] R package v0.6-17.

**4. Technical details for polygenic enrichment analyses, gene prioritization and functional enrichment annotations**

To determine polygenic enrichment between mental disorder phenotypes and suicidality phenotype pairs, shared loci between phenotype pairs were identified using PLACO[12]. PLACO examines one gene at a time with two sets of Z-statistics as input and proceeds by dividing the composite null hypothesis of pleiotropy into three sub-null scenarios: (i) H00: the gene is not associated neither of the two disorders. (ii) H10: the gene is associated with the first disorder but not the second. (iii) H01: the gene is not associated with the first disorder but the second. The alternative hypothesis (H11) is that the gene is related to both disorders, corresponding to pleiotropic association.

Each of the risk loci, determined from functional mapping and annotation (FUMA) (default LD = 0.6), were fine-mapped using CAVIAR[13]. Each The set of causal SNPs were annotated with CADD[14] scores followed by positional gene mapping within ±100 kb. The causal SNPs among all of the PLACO trait-pairs were validated in summary data from both European (suicidality, derived from MTAG results of PGC and MVP) and African ancestry (SITB, MVP)[15].

Summary statistics from the PLACO results were to test for gene-level associations using MAGMA[16], e-MAGMA[17] and h-MAGMA[18] to annotated SNPs derived by PLACO to genes based on SNP location; expression quantitative trait loci eQTL reference dataset and 3D chromatin configuration interaction, respectively. Then, the p-value of each gene was obtained and converted immediately into Z statistic. The direction of Z statistic was determined by the sign of summation of the product of effect sizes and MAFs of all SNPs in each gene. In each suicidality-phenotype pair, genes annotated by both MAGMA and e-MAGMA or h-MAGMA were further functional annotated.

We prioritized all the pleiotropic genes by summary-data-based Mendelian randomization (SMR[6]) analyses, single-cell type from database PanglaoDB and CellMarker 2.0, protein level in UKB OLINK data, OMIM, drug–gene interactions and gene set enrichment analyses for the pleiotropic genes through Metascape and FUMA gen2fun.

In SMR, cis-eQTLs in whole blood from eQTLGen Consortium and 13 brain tissues from GTEx v8 projects[19] (including frontal cortex BA9, hippocampus, nucleus accumbens basal ganglia, cerebellum, caudate basal ganglia, cerebellar hemisphere, hypothalamus, anterior cingulate cortex BA24, cortex, spinal cord cervical c-1, amygdala, putamen basal ganglia and substantia nigra) were used as instrumental variables to link the outcome via an exposure (i.e., gene expression). The analysis was conducted according to default SMR protocols. We further supplied the heterogeneity in dependent instruments (HEIDI) analyses to support inference by reducing the likelihood that linkage disequilibrium affected the SMR findings.

Next, we search these genes in single-cell type database PanglaoDB and CellMarker 2.0 to find brain cell type-specific (neuron, astrocyte, microglia, and oligodendroglia, etc.) expressed genes.

Ordered logistic regression analyses were performed using the R-implemented 'polr' function on OLINK protein level with suicidality, with age, sex, education, and TDI as covariables. In Metascape and FUMA gen2fun, minimum overlapping genes with gene-sets were 3, and maximum FDR adjusted P-value for gene set association was less than 0.05. Enrichment analysis was done for WikiPathways, gene ontology (GO), Kyoto Encyclopedia of Genes, Genomes (KEGG), Reactome Gene Sets and GWAS Catalog. The significance threshold of all the above enrichment analyses was set at FDR < 0.05. We then identified protein–protein interaction (PPI) networks constructed using STRING[20]. From this, we extracted the largest connected PPI networks (protein number ≥ 3) and visualized it using Cytoscape[21]. We identified the natural clusters based on the stochastic flow using the Markov Cluster Algorithm (MCL) method, with inflation parameter equaled to 3.

Finally, prioritized genes were selected to conduct gene-set PRS for SA. SNPs were selected within the start and end base positions of each gene with 10kb extension. PRS association analyses with suicidality in UKB by PRS-CS, adjusted for age, sex, education, TDI, loneliness, whether able to confide, risk taking, chronic pain and whether smoke, top 4 genetic PCs, and array types. To test whether our gene-set PRS was significantly associated with suicidality, a permutation test (10,000 times) was used to build null distributions of all estimates and assess the statistical significance of all latent components at once, and the significance level was set as two-tailed test $P < 0.05$.

### 5. Technical details for two-sample Mendelian Randomization and Multivariate Mendelian Randomization

*5.1 Selection of instrument SNPs*

Only datasets generated from European-ancestry samples were included, and no overlapping individuals were identified between Suicidality and mental disorder GWAS. All SNPs were matched to 1000 Genome Project CEU population[7]. Instrumental SNPs were selected from those with GWAS p $<5\times10^{-6}$ and being uncorrelated ($r^2 < 0.01$). 2SMR was then conducted by the inverse variance–weighted (IVW) method (as primary) and three other companion methods (weighted median, MR-Egger regression imbedded in the TwoSampleMR (0.5.11), MendelianRandomisation (0.9.0). Pleiotropy or heterogeneity test were performed with MR-Egger regression, and MR-PRESSO, respectively. The same criterion as univariable MR was adopted in MVMR (0.2)[22] to select instrumental SNPs for each exposure, covariate, or outcome.

*5.2 detailed methods of MVMR*

For MVMR analyses, we constructed instruments using SNPs in each of the GWASs meeting our single-variable MR selection criteria, described previously. To validate SEM results in cohort analysis part, we combined the SNPs from the relevant GWASs: each mental disorder, mediating brain indices and suicidality. Potential instrumental SNPs with palindromic allelic frequencies were excluded to harmonize multiple exposures. We used the MVMR extension of the inverse-variance weighted MR method and MR-Egger method to correct for both measured and unmeasured pleiotropy.

**Supplementary results**

### 1. Demographic distributions of subgroups in sensitivity analyses.

For SI, SA, SD and SA&SD, there are 144,532, 111,722, 102,403 and 112,000 samples in the clinical association regression analyses; and 141,272, 109187, 93,983 and 109,455 samples in the PRS association regression analyses, respectively.

To separate them into male and female groups, there are 63,619, 51,107, 47,858, and 51,296 male samples and 80,913, 60,615, 50,634, and 60,704 female samples in the clinical association regression analyses; and 62,397, 50,116, 42,333, 50,299 male samples and 78,875, 59,071, 26,144, and 59,156 female samples in the PRS association regression analyses, respectively.

### 2. Total and Mediating Effects of mental disorders on suicidality in AoU

For model 1 of European population, mental disorders were associated with higher incidence of suicidality ($\beta = 0.45$, $p < 0.001$) on condition of PRSs (effect on phenotypes: $\beta = 1.008$, $p < 0.001$) and environmental factors (magnitude of indirect effect: $\beta = 0.252$, $p = 0.01$), and SA PRS was nominal associated ($\beta = 0.008$, $p = 0.115$). As for African population, mental disorders were associated with higher incidence of suicidality ($\beta = 0.45$, $p < 0.001$) on condition of PRSs (effect on phenotypes: $\beta = 1.008$, $p < 0.001$) and environmental factors (magnitude of indirect effect: $\beta = 0.252$, $p = 0.01$), and SA PRS was nominal associated ($\beta = 0.008$, $p = 0.115$) (Fig. S4).

#### 3. Prioritization of Candidate pair-specific and all the pleiotropic genes and Characterization of Phenotype and Tissue Specificity

Among the genes which were pair-specific, 16 were identified as novel with suicidal or mental disorder phenotypes. Several key genes like *DYNC1I2, FAM168A, GABBR1, TMEM258, BAG5, FADS1, KLC1* as were previously validated (both verified in TWAS and single-cell annotation) (Table S15). Moreover, TWAS analyses showed that expression of 6 novel genes were associated with 12 mental disorders in whole blood especially, and 13 brain regions, including *AMT, MED19, TMEM106A, BTN3A3, BAG5* and *KLC1* (Table S16). Further in cell type specificity, 41 genes showed enrichment in multiple sub-types of neuronal cells, astrocytes, glial cells, microglia and others in human or mouse brain-specific tissues (Table S15). We did not find positive association between protein level of pair- specific genes analysis with suicidality in UKB population.

Tissue specificity analysis for all the pleiotropic genes confirmed the tissue specificity not only in brain tissues but also in muscle-skeletal, heart tissue, pancreas, blood and liver (Fig S8).

#### 4. Enriched biological pathways and shared risk factors between suicidality and mental disorders of all the pleiotropic genes

Enrichment analyses determine 125 over-represented pathways among 136 multi-shared genes, including 35 Gene Ontology (GO) terms, 1 Kyoto Encyclopedia of Genes and Genomes (KEGG) pathway and 89 Reactome Gene Sets. The same with multi-shared genes, these enriched pathways are mainly involved in regulation of immune system process and

DNA, nucleosome and chromatin organization. For example, structural constituent of chromatin and nucleosome binding in molecular functions; chromatin remodeling and protein DNA complex subunit organization in cellular components; nucleosome assembly and organization, and chromatin organization and remodeling in biological process are enriched in 4 trait pairs. Additionally, these shared genes were mainly enriched in common mental disorders, brain morphology, intelligence and Sleep duration based on the GWAS catalog (Fig. S9 and Table S21). PPI network analysis (MCL clustering, inflation parameter: 3) showed that 26 sets of 136 multi-shared proteins form a tightly interconnected network (Fig. S9 and Table S22). Fourthly, gene-set PRSs of SA showed significant positive association with suicidality against randomly selected SNPs in UKB ($P$ < 4.17 × 10-3) (Figure S11), indicating their high polygenic effect.

### Supplementary Figures

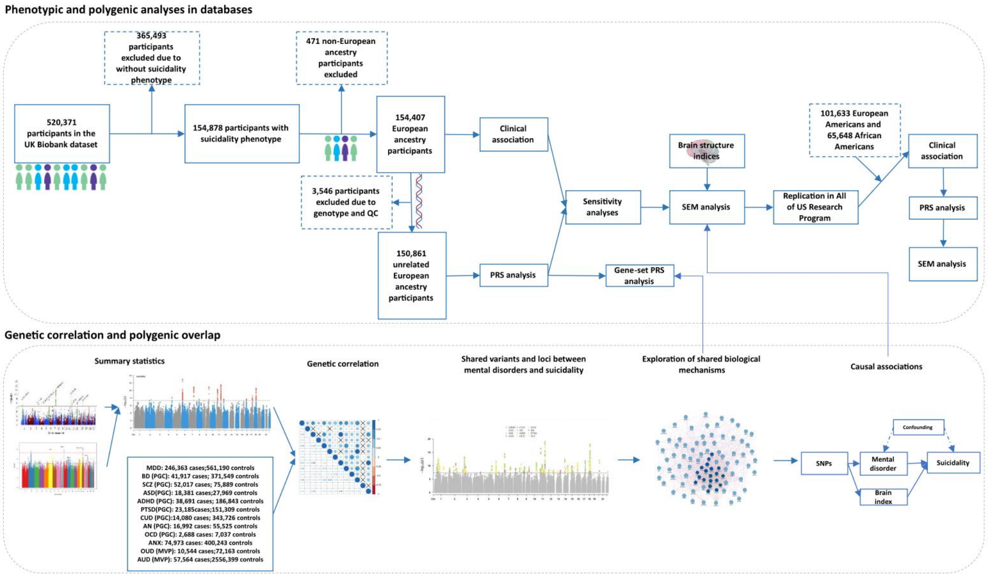


Figure S1: overall analytical pipeline.

Notes: We conducted the comprehensive association and pleiotropic analysis between 12 mental disorders and suicidality from cohort and summary statistics perspectives. PRS: polygenic risk score; SI: suicidal ideation; SA: suicidal attempts; SD: suicidal death; SCZ: schizophrenia; BD: bipolar disorder; MDD: major depressive disorder; ANX: anxiety disorder; OCD: obsessive- compulsive disorder; PTSD: posttraumatic stress disorder; ED: eating disorder; ASD: autism spectrum disorder; ADHD: Attention Deficit Hyperactivity Disorder; AUD: alcohol use disorder; CUD: cannabis use disorder; OUD: opioids use disorder.

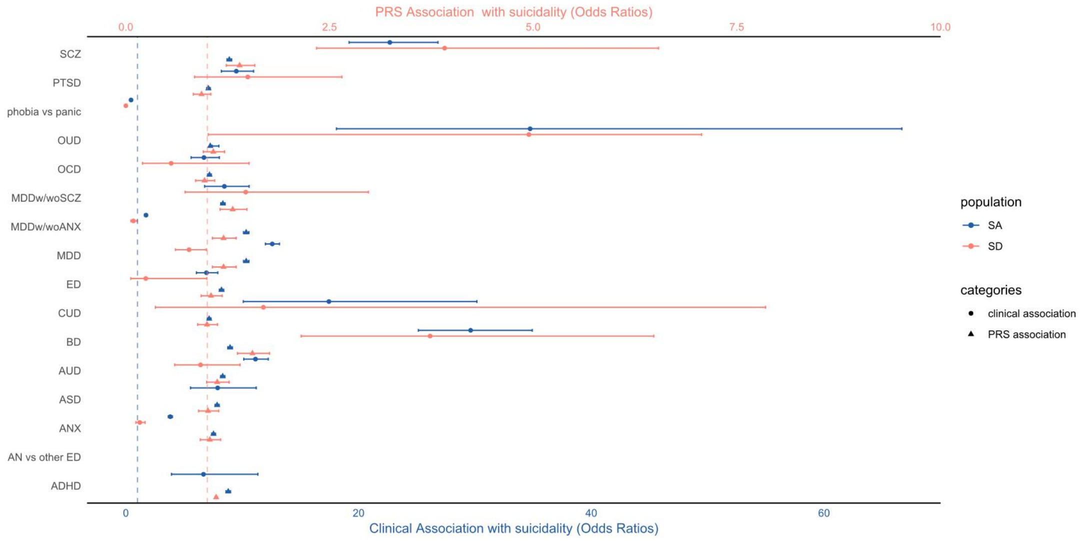


Figure S2:

**Notes:** Associations with SA and SD among various mental disorders and sub-phenotypes. Blue indicates associations with suicide attempts (SA), while red represents associations with suicide deaths (SD). Both blue and red dashed lines indicate an odds ratio (OR) of 1. Circles represent clinical associations, and triangles denote PRS associations. PRS: polygenic risk score; SA: suicidal attempts; SD: suicidal death; SCZ: schizophrenia; BD: bipolar disorder; MDD: major depressive disorder; ANX: anxiety disorder; OCD: obsessive- compulsive disorder; PTSD: posttraumatic stress disorder; ED: eating disorder; ASD: autism spectrum disorder; ADHD: Attention Deficit Hyperactivity Disorder; AUD: alcohol use disorder; CUD: cannabis use disorder; OUD: opioids use disorder; MDDw/woSCZ: MDD with versus without psychosis symptoms; MDDw/woANX: MDD with versus without anxious symptoms.

Figure S3:

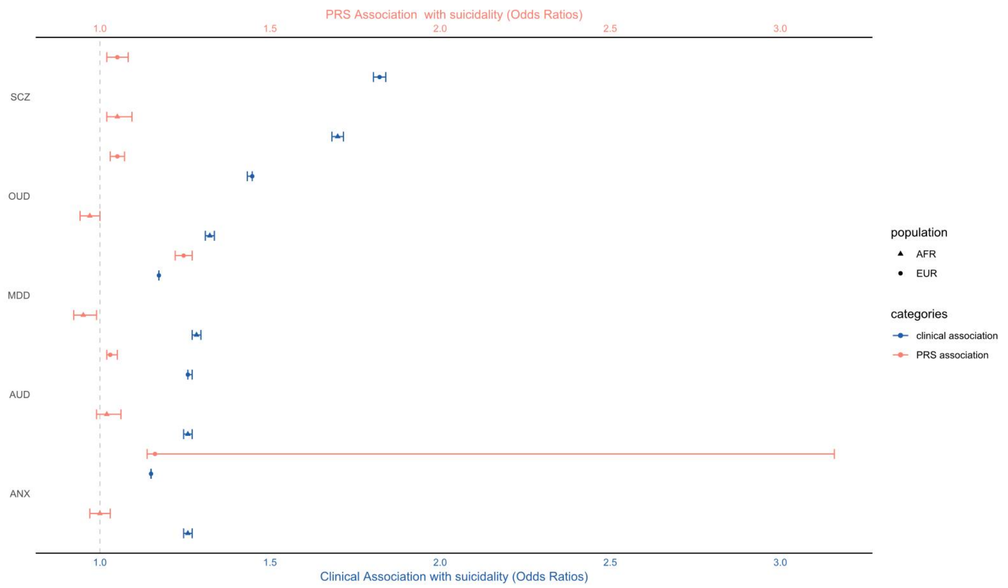


Notes: Clinical and PRS association between suicidality and mental disorders in AoU European and African population. Blue indicates clinical association, while red represents associations with PRS association. Red dashed line indicates an odds ratio (OR) of 1. Circles represent European population, and triangles denote African population. PRS: polygenic risk score; SCZ: schizophrenia;MDD: major depressive disorder; ANX: anxiety disorder; AUD: alcohol use disorder; OUD: opioids use disorder.

Figure S4:

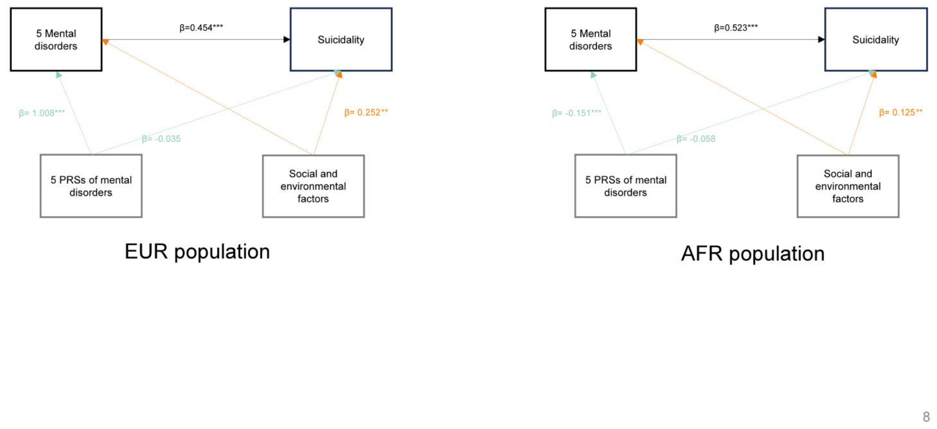


Notes: Structural equational models for 5 mental disorder, PRS, social environmental factors and suicidality in All of Us. Lifetime smoking, multi chronic pain, loneliness, confide, risk-taking, educational status, deprivation index, and BMI are as latent variables for social and environmental factors. *P < 0.05, **P < 0.01, ***P < 0.001.

Figure S5:

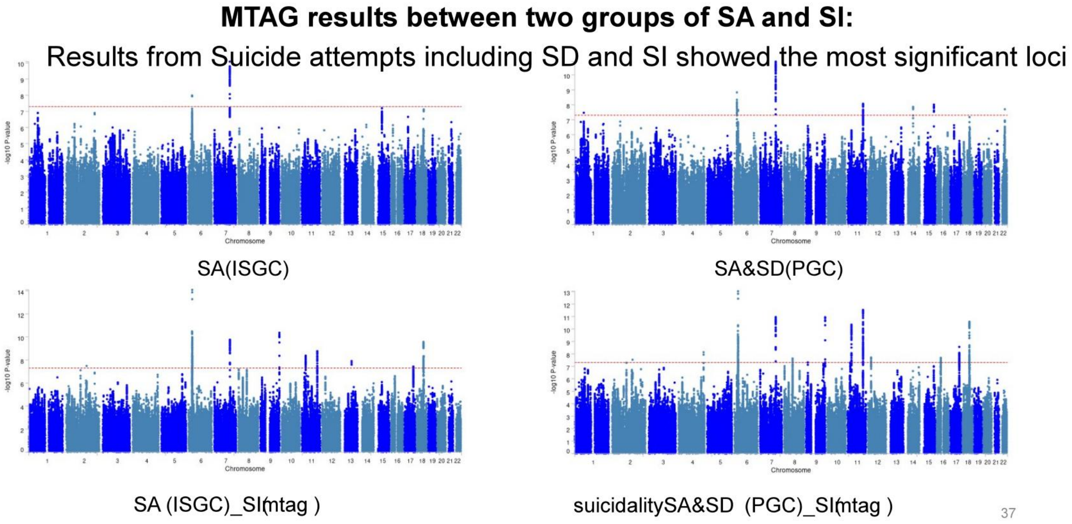


Notes: MTAG results for both SA derived from PGC and SI derived from MVP increases significant loci into 16 SNPs in 13 loci.

Figure S6

### Locuszoom results of newly conducted suicidality GWAS summary data

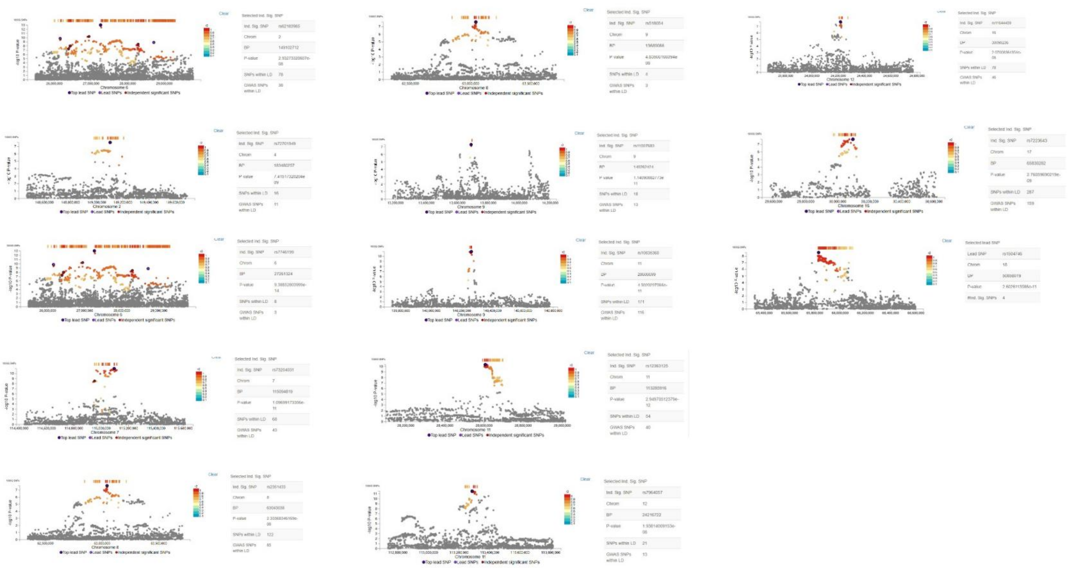

Notes: Locus zoom plots of 13 risk loci of Suicidality.

Figure S7

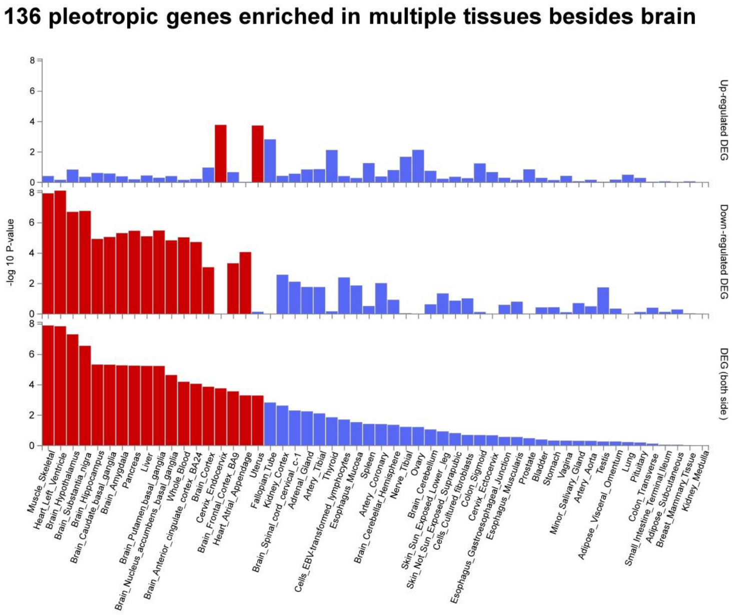


Notes: 136 pleotropic genes enriched in multiple tissues including brain regions and other tissues. Red bars indicate significant differential expression genes (DEG) from up-regulated, down-regulated and both sides.

Figure S8

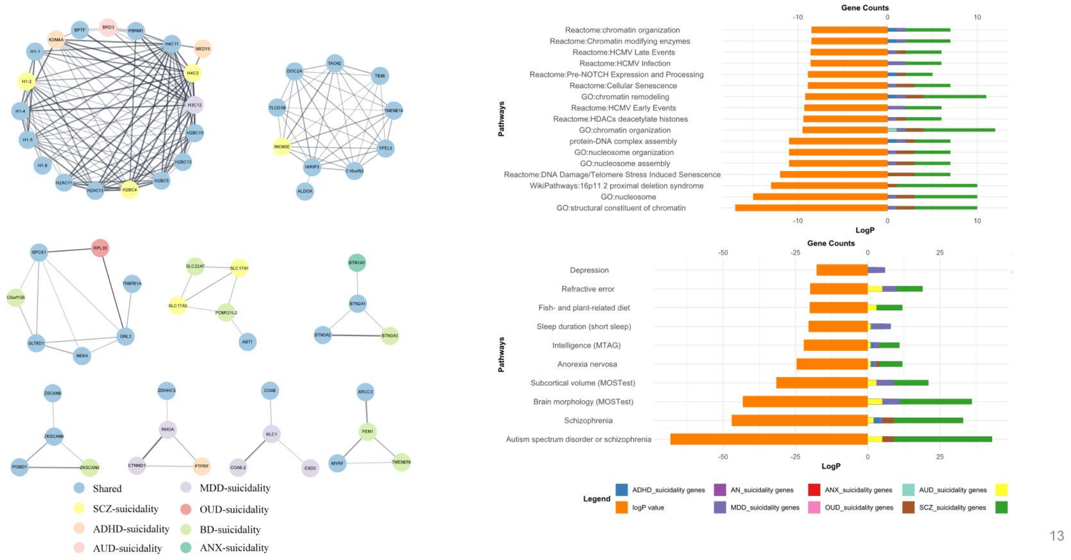


Notes: 136 pleiotropic genes enriched in process of chromosome structure, regulation of immune process, and in mental disorder related traits and brain structures, and 9 PPI network. Different colors represent whether genes are multi-shared, or trait-specific.

Figure S9

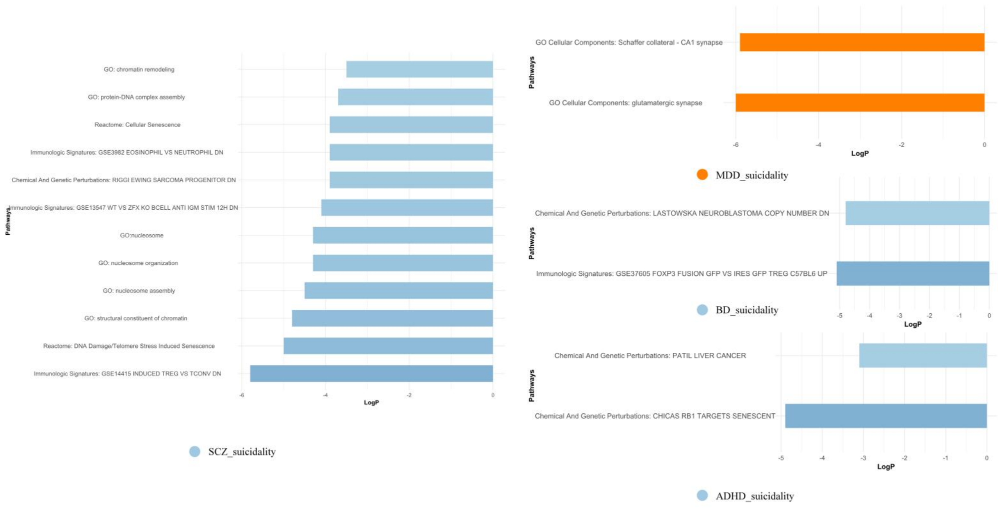


Notes: Pair-specific genes enrichment of SCZ-suicidality, MDD-suicidality, BD-suicidality and ADHD-suicidality pairs.

Figure S10

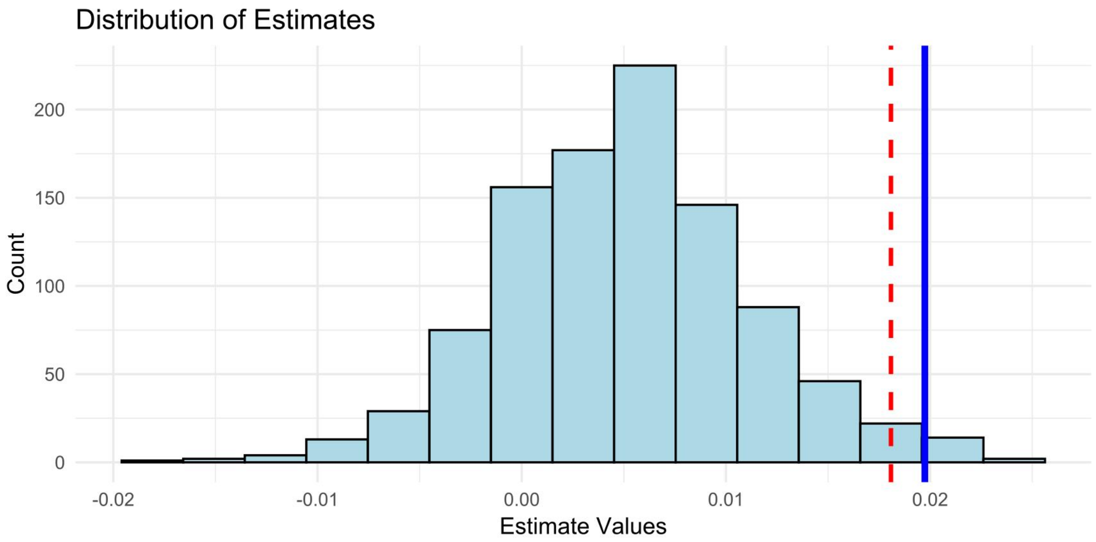


Notes: Gene-set PRS of SA derived from 136 pleiotropic genes showed significantly higher association with suicidality than randomly selected gene-sets.

Figure S11

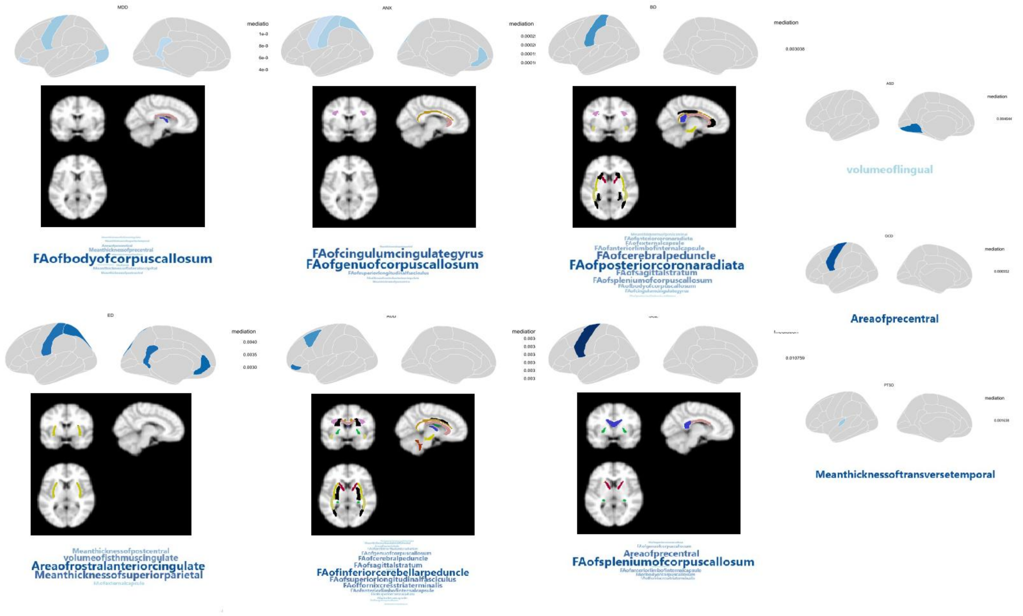


Notes: Mediation effects of brain-related indices on each mental disorder to suicidality. For each mental disorder- suicidality pair, the upper side shows significant mediated structural indices; while the lower side shows white matter integrity in different regions. The font size increases as the effect increases. SCZ: schizophrenia; BD: bipolar disorder; MDD: major depressive disorder; ANX: anxiety disorder; OCD: obsessive- compulsive disorder; PTSD: posttraumatic stress disorder; ED: eating disorder; ASD: autism spectrum disorder; AUD: alcohol use disorder.
